# Usefulness of the smartphone app-based smoking cessation program for conventional cigarettes, heated tobacco products, and dual use: A retrospective study

**DOI:** 10.1101/2022.07.13.22277567

**Authors:** Yuko Noda, Ryuhei So, Misaki Sonoda, Takahiro Tabuchi, Akihiro Nomura

**Author notes:** **Corresponding author:** Akihiro Nomura, MD, PhD, Department of Biomedical Informatics, CureApp Institute, 4136-1 Azayakozawa, Nagakura, Karuizawa-machi, Kitasaku-gun, Nagano 389-0111 Japan.

## Abstract

**Background:** Heated tobacco products (HTPs) are widespread in Japan, and smoking cessation of such products have become an important issue owing to the dissemination of the harmful effects of HTPs. The efficacy of online digital therapy has been reported in smoking cessation treatment; however, we have limited evidence of online smoking cessation program for HTP users. In this study, we evaluate the usefulness of the “ascure” program for people using HTPs (exclusive HTP use or dual use of HTP and cigarettes) compared with exclusive cigarette users.

**Methods:** This was a retrospective study. We recruited adult smokers participating in the ascure online smoking cessation program in Japan from June 2019 to February 2021. The primary outcome was the chemically validated continuous abstinence rate (CAR) at weeks 21–24 using salivary cotinine testing. We also assessed CAR 9–12 and program adherence.

**Findings:** We analyzed data from 2952 participants (52% for cigarette group, 35% for HTP group, and 13% for dual-use group), the mean age was 43·4 ± 10·8 years, 17% were women. Exclusive HTP user were more likely to stop tobacco use than exclusive cigarette smokers in CAR 21–24 (52.6% for cigarettes vs. 64.8% for HTP; OR: 1·17; 95% CI: 1·12–1·22; p < .001).

**Interpretation:** Exclusive HTP users had higher CARs and adherence compared with exclusive cigarette users, indicating a higher affinity for the ascure online smoking cessation program. This program might be a useful smoking cessation option for HTP users as well as cigarette smokers.

**Funding:** CureApp, Inc.

**Research in context:** *Evidence before this study:* Heated tobacco products have been widespread in Japan recently, and smoking cessation of these tobacco products became an important issue due to the gradual revealing of HTPs’ harmful effects on health. Besides, the efficacy of new online digital therapy has been reported in smoking cessation treatment. We searched PubMed on Dec 14, 2021, for publications on the effects of Heated tobacco on health and its relation to smoking cessation, using the terms “Heated tobacco products”, “smoking cessation” and “ digital therapy “ with no restrictions on publication date. We excluded studies that reported specifically on electronic cigarettes. We found evidence about the harmful effect on the health of using HTPs, and the status and issues of smoking cessation, a moreover new attempt at smoking cessation using digital therapy. However, the efficacy of smoking cessation treatment was mostly limited to conventional cigarettes especially digital therapy we found little evidence regarding smoking cessation by tobacco products such as HTPs.

*Added value of this study:* In this study, we evaluated the usefulness of the ascure online smoking cessation program by tobacco products with a chemically validated index. The continuous abstinence rates (CARs) 21-24 was higher in the exclusive HTP group than the exclusive cigarettes group. Moreover, there was no significant difference between the exclusive cigarettes and the dual-use groups.

*Implications of all the available evidence:* The number of HTP users is currently increasing in Japan. In addition, online medical treatment is becoming more widespread and is expected to solve medical issues. The results of this study indicating a higher affinity for the ascure online smoking cessation program. This program might be a useful smoking cessation option for HTP users.

## Introduction

Smoking causes many diseases, most notably cardiovascular disease, chronic obstructive pulmonary disease, and cancer. Smoking is one of the major risk factors for adult mortality from non-communicable diseases in Japan.^1,2^ Despite the government’s smoking cessation efforts such as increasing tobacco taxes and promoting the prevention of passive smoking, the national adult smoking rate remains high in Japan: 27·1% for men and 7·6% for women as of 2019.^3,4^ Tobacco companies have also aggressively promoted heated tobacco as an alternative product to conventional cigarettes. Since Philip Morris first launched the world’s first heated tobacco product (HTP)—iQOS—in Japan and Italy in 2014, 27·2% of male and 25·2% of female smokers in Japan now use the product.^4^ In addition, 6·9% of men and 4·8% of women reported dual use of conventional cigarettes and HTPs.^4^ Since HTPs have similar hazardous effects, such as cardiovascular diseases, as traditional cigarettes,^5,6^ it is necessary to encourage people to quit using both HTPs and conventional cigarettes.^7,8^

A standard smoking cessation program is provided at outpatient clinics in Japan for people with nicotine dependence to quit using tobacco. Moreover, HTP users have been eligible for smoking cessation treatment under general health insurance since 2020. A standard smoking cessation program consists of physicians’ counseling and pharmacotherapy using varenicline or a nicotine patch for 12 weeks in-person or through telemedicine.^9^ However, the program’s completion rate is low (36%) because most patients receiving smoking cessation treatment are of working age and are too busy to visit the outpatient clinics.^10^ In addition, the continuous abstinence rate (CAR) drops significantly after completing the 12-week program.^11^

To reduce program withdrawal and to support long-term smoking cessation, CureApp, Inc. released the ascure online smoking cessation program.^12–14^ The program provides complete face-to-face telemedicine services using pharmacotherapy for physical dependence and behavioral therapy for psychological dependence. Interventions that combine pharmacotherapy and behavioral therapy increase smoking cessation success.^15,16^ We previously reported the favorable smoking cessation success rates of the ascure online smoking cessation program for participants who used traditional cigarettes. However, there is no evidence of usefulness regarding the ascure online smoking cessation program for HTP users.

Hence, we performed a retrospective analysis to evaluate the usefulness of the ascure online smoking cessation program for people using HTPs (exclusive HTP or dual use of HTP and cigarettes) compared with exclusive cigarette users.

## Methods

### Study design

This was a retrospective study to evaluate the usefulness of the ascure online smoking cessation program for HTP users. In brief, we divided the program’s participants into three groups based on their tobacco product use (exclusive cigarettes group, exclusive HTP group, or dual use of both cigarette and HTP group). Then, we compared the smoking cessation success rates of the exclusive cigarette group (as reference) with the exclusive HTP or dual-use groups. The primary endpoint was the biochemically validated CAR at weeks 21 to 24 (CAR 21–24). This study was conducted in accordance with the Declaration of Helsinki, the Ethical Guidelines for Medical and Biological Research Involving Human Subjects, and all other applicable laws and guidelines in Japan. The study protocol was approved by the Institutional Review Board of Kanazawa University, Japan (no. 2021-184 [113839]). Since anonymized information was used for the analysis, no written consent was obtained. However, prior to using the ascure smartphone app, participants were clearly informed that the app’s data would be used for research; only those who consented to this could use the app.

### Participants

We recruited adult smokers who participate in the ascure online smoking cessation program in Japan from June 2019 to February 2021. We included participants who met all of the following criteria: 1) enrolled in the affiliated Japanese health insurance association; 2) willing to quit using tobacco immediately; 3) could use a smartphone (OS: Android^®^ 5·0 or higher, iPhone^®^ 10·0 or higher); and 4) agreed to participate in the smoking cessation program in the app. We excluded participants who had severe mental illness or difficulty continuing the entire program. For analysis, we also excluded participants without sufficient baseline information.

### Outcomes

The primary outcome was CAR 21–24. We defined smoking cessation success as self-reported successful smoking cessation for the past month during the interview and confirmed this by salivary cotinine testing as a chemical validation.^17^ We used the Nicotine Dependence Cognition Scale (NDCS), which indicates the severity of nicotine dependence and cognitive impairment in smokers.^14^ The secondary outcomes were CAR at weeks 9 to 12 (CAR 9–12), the impact of tobacco products on the progress of quitting smoking, and the program adherence rates.

### Ascure online smoking cessation program

Figure 1 provides an overview of the ascure online smoking cessation program. Participants can receive the on-demand video tutorial through the app anywhere, and at any time, and there are six to eight online-mentoring smoking cessation counseling sessions by experienced nurses and pharmacists. All online counselors completed in-house training before conducting the interviews, and the training included instructions on smoking cessation as well as skills on how to explain the appropriate use of the app. In addition, the participants can be concurrently administered over-the-counter nicotine patches or nicotine gum for 8 weeks.

**Figure 1.**
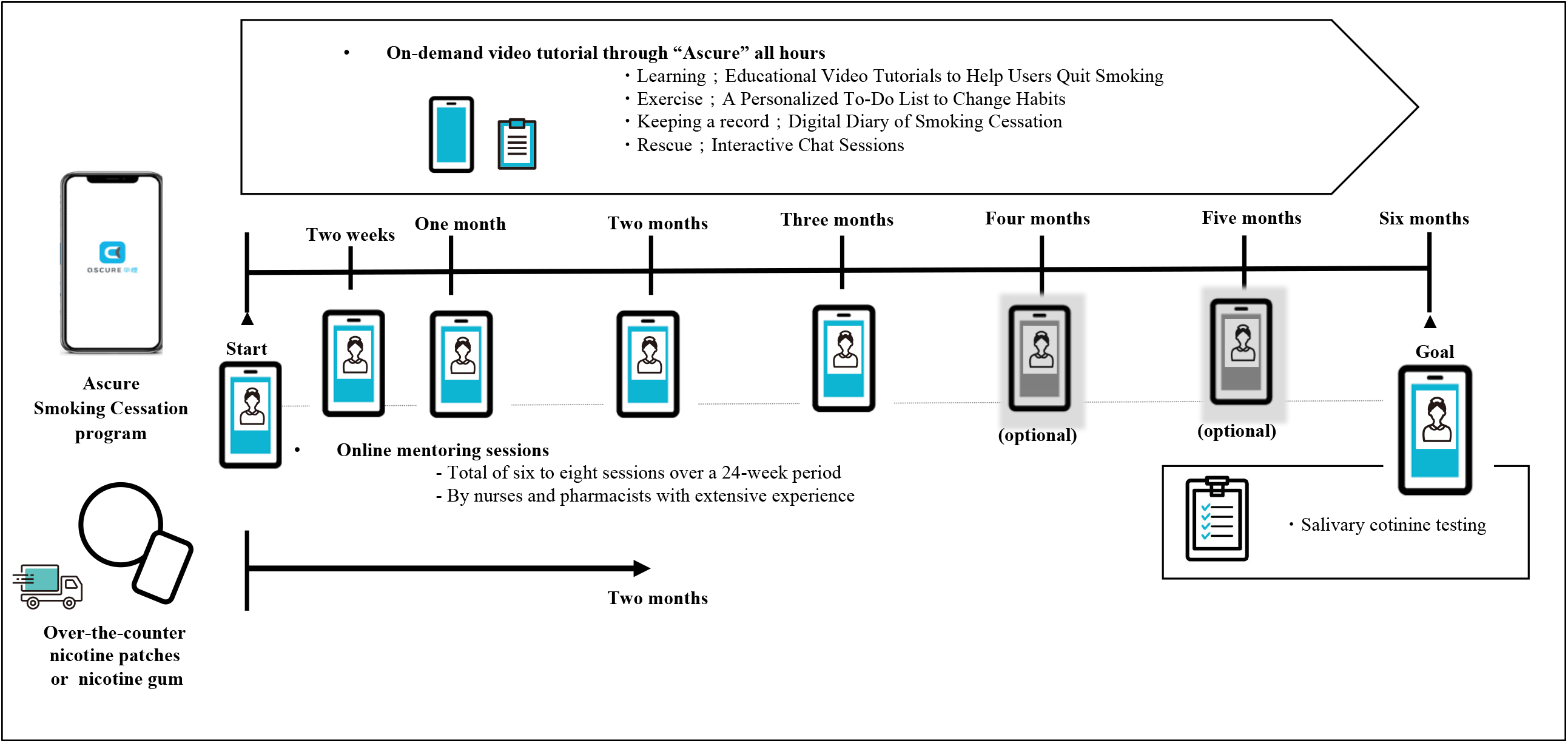
Overview of the ascure online smoking cessation program for smoking cessation. Participants can receive the on-demand video tutorial through the app anywhere, and at any time, and there are six to eight online-mentoring smoking cessation counseling sessions by experienced professionals. The app consists of four elements: 1) Learning; educational video tutorials to help users quit using tobacco, 2) Exercise; a personalized to-do list to change habits, 3) Keeping a record; a digital diary of smoking cessation, and 4) Rescue; interactive chat sessions for relief from cravings or withdrawal symptoms. In addition, the participants can be concurrently administered over-the-counter nicotine patches or nicotine gum for 8 weeks. In the end, participants were taken the chemically validated continuous abstinence rate (CAR) at six months using salivary cotinine testing.

**Figure 2.**
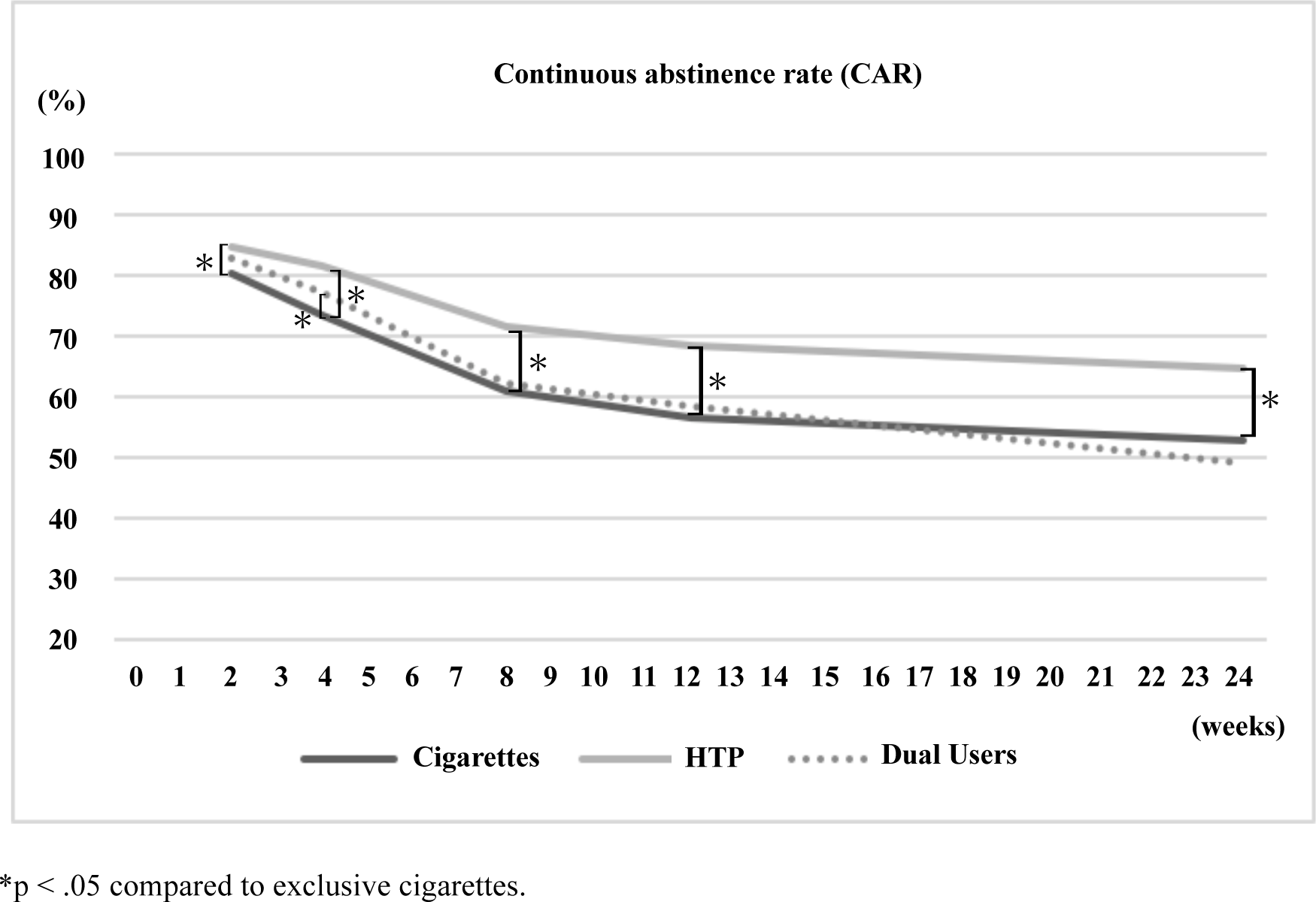
CARs among the three groups. CAR, continuous abstinence rate. The biochemically validated CAR 21–24 was 52·6% in the exclusive cigarettes group, 64·8% in the exclusive HTP group, and 48·7% in the dual-use group. Compared with the exclusive cigarettes group, the exclusive HTP group had significantly higher CAR 21–24, whereas there was no significant difference between the exclusive cigarettes and the dual-use groups.

The ascure smartphone app was released by CureApp, Inc. (Tokyo, Japan). Participants can download the app to their own smartphones and start using it by entering a unique passcode individually issued by CureApp. After installing the app, participants proceed to the appointment for the first interview if they agree to the Personal Information Protection Regulations, including consent to the anonymized information data analysis. The app consists of four elements: 1) educational video tutorials to enhance the understanding of nicotine dependence, 2) a personalized to-do list for behavior change, 3) a digital diary for record-keeping, and 4) interactive chat sessions for relief from cravings or withdrawal symptoms.

Details of the app have been provided elsewhere.^17^

### Data collection

We collected each participant’s information, including tobacco product types, through the app and counseling sessions. The counselors checked the success or failure of smoking cessation at each interview based on participants’ self-reports. Data acquired by the app included age, sex, number of cigarettes smoked per day, years of tobacco use, and motivation to quit using tobacco.

### Statistical analysis

Baseline characteristics were described using means (standard deviation) for continuous variables or numbers (proportion in %) for categorical variables. We analyzed the primary outcome using inverse probability weights (IPW) against tobacco product type estimated by multinomial propensity scores based on 5000 regression trees and the average treatment effect on the treated. The propensity model included age, sex, Brinkman Index, cigarettes per day, years of tobacco use, number of smoking cessation attempts before the study, and NDCS as covariates.^18,19^ We presented both before and after IPW maximum standardized differences of each baseline characteristic. We defined the maximum standardized difference of 0·2 or higher as an indication of imbalance.^18,19^ All analyses were performed using R software version 3·6·3 (R Foundation for Statistical Computing, Vienna, Austria), with p-values < ·05 deemed significant.

## Results

### Baseline characteristics

A total of 3478 individuals participated in the ascure online smoking cessation program. Of those, we excluded 448 participants without tobacco product type information and 78 without sufficient baseline details. Thus, we enrolled 2952 participants (1524 for the exclusive cigarette group, 1038 for the exclusive HTP group, and 390 for the dual-use group) for further analyses.

Table 1 shows the participants’ characteristics at baseline before IPW. The mean age was 43·4 ± 10·8 years, 17% were women, and the mean years of tobacco use was 22 years. We used IPW to minimize the differences of baseline characteristics among the groups. The characteristics were well-balanced after applying IPW: all maximum standardized differences among the groups were ≤ ·05 (Table 1).

**Table 1.** Participants’ clinical characteristics at baseline.

|  | Total | Exclusive<br>cigarettes | Exclusive<br>HTP | Dual use | Maximum<br>standardized<br>difference * |  |
| --- | --- | --- | --- | --- | --- | --- |
|  | N = 2952 | n = 1524 | n = 1038 | n = 390 | Before | After |
| Age (years), mean (SD) | 43.4 (10.8) | 44.6 (11.0) | 42.2 (10.1) | 41.4 (11.0) | 0.30 | 0.05 |
| Female sex, n (%) | 513 (17.4) | 286 (18.8) | 178 (17.1) | 49 (12.6) | 0.16 | 0.05 |
| Brinkman Index, mean (SD) | 392 (290) | 411 (298) | 376 (285) | 364 (261) | 0.16 | 0.00 |
| Cigarettes per day, mean (SD) | 17.5 (8.9) | 17.2 (7.4) | 17.9 (11.2) | 17.4 (6.7) | 0.10 | 0.04 |
| Years of tobacco use, mean (SD) | 21.6 (10.8) | 22.8 (11.2) | 20.4 (10.0) | 20.1 (11.0) | 0.24 | 0.01 |
| Number of smoking cessation attempts<br>before the study, mean (SD) | 1.7 (2.1) | 1.7 (2.2) | 1.6 (1.9) | 1.6 (2.0) | 0.07 | 0.05 |
| NDCS**, mean (SD) | 11.8 (3.5) | 11.9 (3.6) | 11.7 (3.5) | 11.5 (3.5) | 0.10 | 0.02 |

\*Maximum standardized mean difference before and after inverse probability weighting.
Abbreviations: NDCS, Nicotine Dependence Cognition Scale; SD, standard deviation.

### Overall smoking cessation success rate using the ascure online smoking cessation program

First, we evaluated the overall success rates of the ascure online smoking cessation program. The overall biochemically validated and CAR 9–12 was 61·0% (95% CI: 59·2–62·8), and CAR 21–24 was 56·4% (95% CI: 54·6–58·2).

### Primary endpoint

The biochemically validated CAR 21–24 was 52·6% (95% CI: 50·1–55·1) in the exclusive cigarettes group, 64·8% (95% CI: 61·9–67·7) in the exclusive HTP group, and 48·7% (95% CI: 43·7–53·7) in the dual-use group (Table 2). Compared with the exclusive cigarettes group, the exclusive HTP group had significantly higher CAR 21–24 (OR: 1·17; 95% CI: 1·12–1·22; p < .001), whereas there was no significant difference between the exclusive cigarettes and the dual-use groups (OR: 0·99; 95% CI: 0·93– 1·05; p = ·77).

**Table 2.** Continuous abstinence rates by tobacco products.

|  | Exclusive<br>cigarettes | Exclusive<br>HTP | Dual use | OR for<br>cigarettes (HTP<br>vs Cigarettes) | P-value<br>(HTP vs<br>Cigarettes) | OR for<br>cigarettes (Dual<br>vs Cigarettes) | P-value<br>(Dual vs<br>Cigarettes) |
| --- | --- | --- | --- | --- | --- | --- | --- |
| **Continuous abstinence rates** |  |  |  |  |  |  |  |
| CAR 9–12 | 56.7%<br>[54.2–59.2] | 68.3%<br>[65.5–71.1] | 58.2%<br>[53.3–63.1] | 1.15<br>[1.10–1.19] | < .001 | 1.05<br>[1.00–1.12] | 0.072 |
| CAR 21–24 | 52.6%<br>[50.1–55.1] | 64.8%<br>[61.9–67.7] | 48.7%<br>[43.7–53.7] | 1.17<br>[1.12–1.22] | < .001 | 0.99<br>[0.93–1.05] | 0.77 |

Values are reported with 95% CI. P-values were calculated using an inverse probability weighted dataset.

### Secondary endpoint

CAR 9–12 was 56·7% (95% CI: 54·2–59·2) in the exclusive cigarette group, 68·3% (95% CI: 65·5–71·1) in the exclusive HTP group, and 58·2% (95% CI: 53·3–63·1) in the dual-use group. Compared with the exclusive cigarettes group, the exclusive HTP group also showed significantly higher CAR 9–12 (OR: 1·15; 95% CI: 1·10–1·19; p < .001) (Table 2). In contrast, there was again no significant difference between the exclusive cigarettes group and the dual-use group (OR: 1·05; 95% CI: 1·00–1·12; p = ·072). Figure 3 shows the proportions of successes, failures, and dropouts of smoking cessation in CARs. The program adherence rate at week 24 was 70·7% overall: 68·4% in the exclusive cigarette group, 75·0% in the exclusive HTP group, and 67·9% in the dual-use group. CAR 21–24 among people who completed the program was 76·8% in the exclusive cigarettes group, 86·4% in the exclusive HTP group, and 71·7% in the dual-use group.

**Figure 3.**
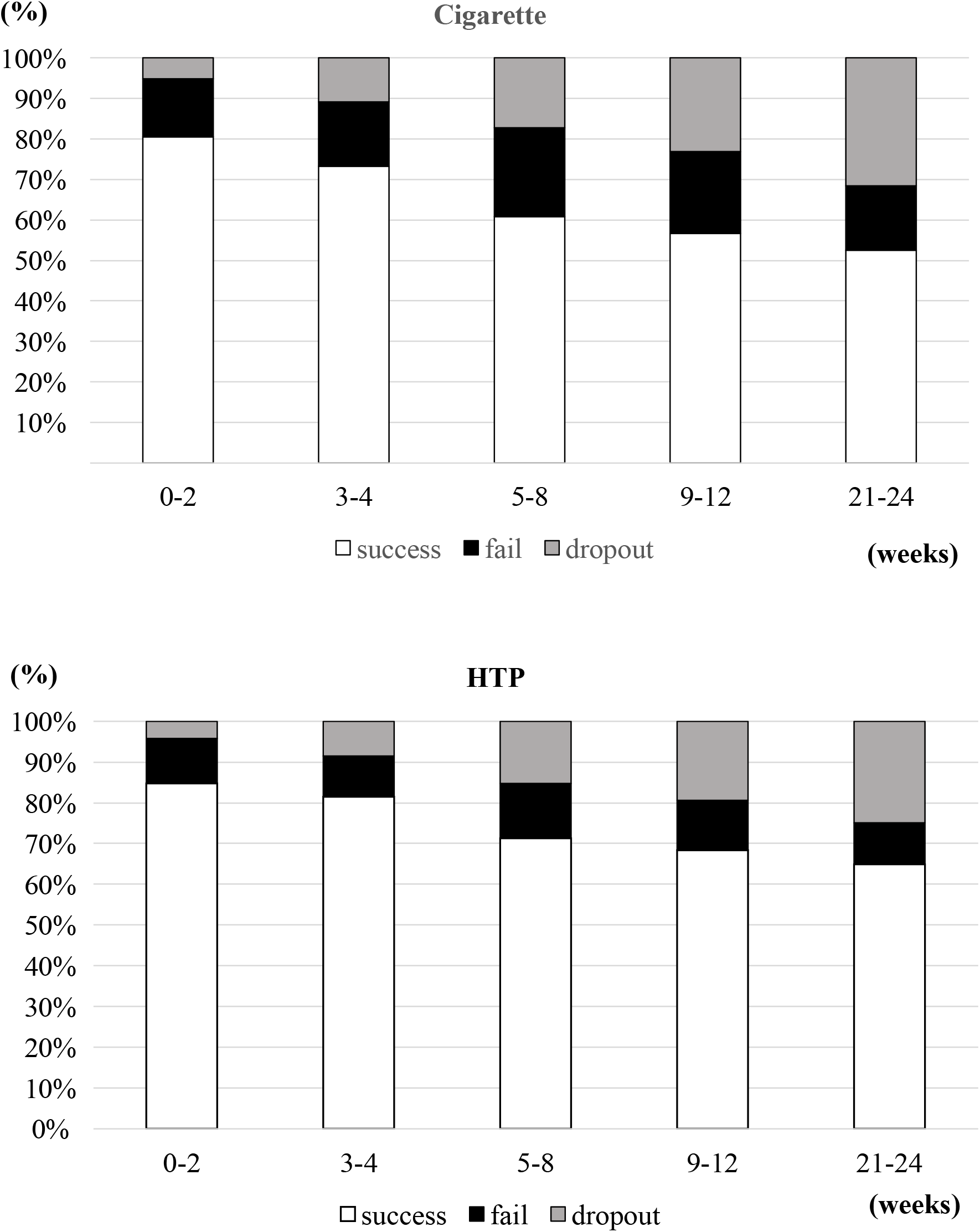
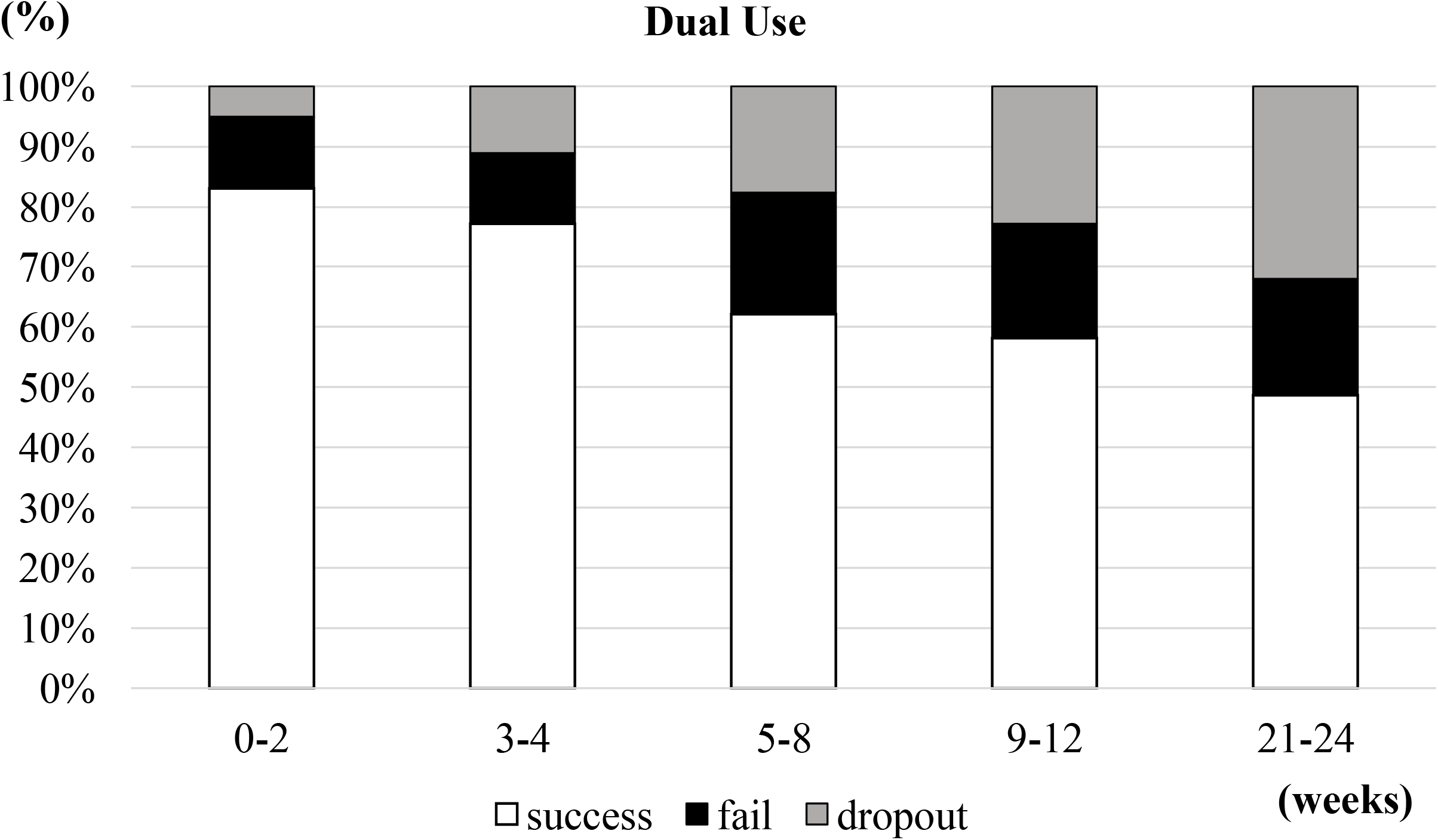
Proportions of successes, failures, and dropouts of smoking cessation in CARs by tobacco products (Cigarette, HTP, Dual Use). The program adherence rate at week 24 was 68·4% in the exclusive cigarette group, 75·0% in the exclusive HTP group, and 67·9% in the dual-use group.

## Discussion

In this study, we evaluated the usefulness of the ascure online smoking cessation program for people using HTPs compared to exclusive cigarette users. We found that there was a significantly higher CAR 9– 12 and CAR 21–24 in the exclusive HTP group compared to the exclusive cigarettes group, whereas there was no significant difference in CAR 9–12 and CAR 21–24 between the exclusive cigarettes group and the dual-use group. In addition, the program adherence rates were favorable in all groups. Specifically, the HTP users had a higher ascure online smoking cessation program adherence rate compared with the excursive cigarettes users.

This study had several findings. First, the biochemically validated CAR 9–12 and CAR 21–24 were significantly higher in the exclusive HTP group compared to the exclusive cigarettes group. There are several possible reasons for the favorable result. First, many people might use HTPs as assistance for smoking cessation,^20^ and these individuals tend to be highly motivated to quit using tobacco. Second, previous reports have shown that more HTP users meet the lower nicotine dependence and patient background conditions that are predictors of smoking cessation success compared to cigarette users.^18,21^ Moreover, because HTPs was not available prior to the launch of HTPs in 2014, the exclusive HTP users once had experienced successful replacement from conventional cigarettes to HTPs completely in average of 20 years of tobacco use experience. Past success in quitting smoking also contributes to future success in quitting smoking.^21^ Furthermore, the higher program adherence rate could contribute to the increase in CAR.^9,17^ The higher adherence rate in the exclusive HTP users compared to that in the exclusive cigarettes users might be attributed to favorable CAR. Nomura et al. previously reported the efficacy of a telemedicine smoking cessation program for nicotine-dependent people who used HTPs over an 8-week telemedicine program provided by primary physicians and a 10-month follow-up period (sending surveys and smoking cessation advice via the app).^22^ They found that exclusive HTP users had significantly higher CAR 9–24 than the exclusive cigarettes group (53·8% for cigarettes vs. 67·0% for HTP). These reports are consistent with the present results, and they suggest the efficacy of the online smoking cessation treatment for exclusive HTP users.

Second, the biochemically validated CAR 9–12 and CAR 21–24 were not significantly different between the dual-use group and the exclusive cigarettes group. Advantages and disadvantages could exist regarding dual users’ smoking cessation. As an advantage, using HTPs could assist with smoking cessation.^20^ Dual users are already in the process of implementing smoking cessation practices. In addition, clinical characteristics of dual users are more likely to be male compared to conventional cigarette users, that meet the predictors of successful smoking cessation and could be the advantage of smoking cessation.^21^ As a disadvantage, dual users tend to feel inadequate for exclusive HTP use,^23^ which might inhibit smoking cessation success and full transition to HTPs. Prior studies have reported lower smoking cessation rates for dual users as compared to their counerparts.^22^ Another report showed no difference in smoking cessation behavior between dual users and cigarette users.^24^ Thus, the present results might be the consequence of the offsetting advantages and disadvantages among dual users.

Several factors like lower age and lower recognition about the degree of harmfulness are known to increase re-smoking rates.^25^ Dual users tend to have a larger CAR drop from week 12 to 24 than the exclusive cigarette group and meet risk factors for re-smoking^24^ Thus, continued observation after the therapeutic intervention is needed, especially for dual users, considering their risk of re-smoking.

Third, the program adherence rates were favorable: 70·7% overall. Specifically, the exclusive HTP users had higher adherence rates compared with the exclusive cigarettes users. Kato et al. previously reported that program adherence was 59·9% at week 24 using the same ascure online smoking cessation program.^17^ Our study had a high proportion of male participants as well as exclusive HTP users, who tend to have a high adherence rate, which likely contributed to the overall improvement in adherence.^9^ Moreover, approximately half of the current smokers in Japan use HTPs because these products might help them quit using tobacco.^20^ Moreover, exclusive HTP users are more likely to report that HTP is helpful for smoking cessation than dual users are.^26^ Exclusive HTP users might have higher motivation and awareness to quit using tobacco than traditional smokers, which might contribute to the high program adherence rate.

The strengths of our study include analyzing data from many participants, specifically, the 1524 people in the exclusive HTPs group. We also used the salivary cotinine test as a chemical validation for the success of smoking cessation. Self-reports of smoking status could be inaccurate, and the salivary cotinine test is known to be a highly sensitive measure to ascertain smoking status.^27^ Measuring nicotine metabolites in saliva is useful in HTP users, too since HTP contains nicotine as well as conventional cigarettes.^28^ Despite these strengths, this study had some limitations. First, we used CAR 21–24 as the primary outcome. Although CAR 9–24 is important for the long-term assessment of smoking cessation,^29^ we could not collect the series of success or failure smoking cessation outcomes from weeks 13 to 20 because of the optional counseling session period. Second, the true success or failure of smoking cessation was unknown for those who dropped out of the program. Although we considered them as smoking cessation failure, some might be misclassified, and they in fact withdrew from the program owing to smoking cessation success.

In conclusion, the exclusive HTP users had higher CARs and adherence rates compared with the exclusive cigarette users, indicating a higher affinity for the ascure online smoking cessation program. This program might be a useful smoking cessation option for HTP users. Whereas, since smoking cessation was not easy among dual users, further research on the complex mechanisms in dual users of smoking cessation is warranted.

## Contributors

YN, AN, RS, and MS designed the analytical strategy and all authors helped to interpret the findings. YN and AN conceived the study. YN performed the data analysis, which was reviewed and interpreted by AN and RS. YN and AN wrote the manuscript, and RS, MS, and TT reviewed and commented on its content. All authors revised the manuscript for critical content and approved its final version. YN and AN had full access to all the data in the study and all authors could access the data on request. AN had final responsibility for the decision to submit for publication.

## Data Availability

The data underlying this study have been prepared under ethical considerations for specific research purposes and are not available to the public.

## Acknowledgments

We thank Naoto Imamachi; software engineer, Naka Katsunori; general manager, Rina Kawagota; product manager, administrative staff, all the participants involved in this program, and medical staff as followed; Seika Irie, Megumi Hayakawa, Tomoko Takahashi, Keiko Ishiba, Meika Taguchi, Ayaka Miyoshi, Riho Nakamura, Yui Enomoto, Momoko Sato, Hayama Noju, Hide Nakamura, Rei Tasaki, Kazuyo Kono, Yui Sugawara, Karen Nakama, Chiaki Kurihara, Saika Sugihara, Haruna Ishizaka, Atsuki Ueno, Rikako Kuroki, Chito Kanda, Misaki Nigawara, Misa Hizaki, Risa Takashio, Kiho Miyoshi. We also thank Editage (www.editage.com) for English language editing.

